# Gender and the associations between HIV and TB-related stigma and mental health outcomes among people with tuberculosis in Botswana

**DOI:** 10.1101/2025.11.14.25340213

**Authors:** Emmanuel Sakyi, Annabelle Wu, Davena Duru, Tamara Jimah, Nakia C. Best, Keneilwe Molebatsi, Chawangwa Modongo, Sanghyuk S. Shin

## Abstract

Tuberculosis and HIV are highly stigmatized, and co-infection can result in intersectional stigma worsening mental health outcomes (depression and anxiety). Meanwhile, stigma may be experienced differently by gender. The study aimed to examine the associations between TB and HIV-related stigma and mental health outcomes, and to assess whether gender moderates these relationships among people newly diagnosed with TB in Botswana. Participants were recruited from 12 health centers in Gaborone. Depression, anxiety, TB-related Stigma and HIV-related Stigma were assessed using PHQ-9, the ZUNG Self-Rating Anxiety Scale, the Van-Rie TB-related Stigma Scale and an HIV-related Stigma Scale, respectively. Association between mental health outcomes and TB or HIV-related stigma was examined using linear regression. Moderation analysis was done by using interaction terms. Among 180 participants with TB, 64 (35.6%) were women and 99 (55.0%) had HIV. The association between depression and stigma (TB-related stigma from patient perspective, TB-related stigma from community perspective, HIV-related stigma) did not reach statistical significance after adjusting for confounders (β = 0.12, p = 0.065; β = 0.03, p = 0.7; β = 0.07, p = 0.4, respectively). Similarly, the regression models for anxiety and stigma (TB-related stigma from patient perspective, TB-related community perspectives, HIV-related stigma) also showed no evidence of association in the final models (β = 0.04, p = 0.7; β = −0.11, p = 0.4; β = 0.24, p = 0.14, respectively). Food insecurity showed a positive association with mental health outcomes whereas male gender was associated with lower depression and anxiety. There was no evidence of gender moderation for the association between mental health outcomes and the various stigma types. We recommend that TB programs should be strengthened to address food insecurity as well as other financial and material needs of people with TB and or HIV.

## Introduction

Tuberculosis (TB) is an infectious disease of the lungs caused by the *Mycobacterium tuberculosis* (Mtb) bacterium [1]. Among infectious diseases, TB is the leading causes of death according to the World Health Organization (WHO). The WHO estimates that TB affects over 10 million people globally, leading to 1.5 million annual deaths [2]. Current data from the World Health Organization also indicate that most TB cases occur in low-and middle-income countries [2,3]. Similarly, HIV remains a major public health concern since its discovery in the 1980s. So far, HIV/AIDS has claimed the lives of over 42 million people worldwide [4]. Despite substantial progress in HIV treatment and prevention efforts, the infection continues to spread. Approximately 40 million people were living with HIV/AIDS at the end of 2023, with about 1.3 million new infections recorded [5]. HIV and Tuberculosis are intricately interconnected. HIV infection leads to a weakened immune system; hence, people living with HIV are more likely to develop active TB and have worse TB outcomes [4,5]. Consequently, TB is the leading cause of death among people living with HIV infection [2].

In spite of medical advancements, social determinants of health such as stigma continue to pose considerable barriers to TB/HIV care and health outcomes [6]. Stigma is a process by which people are discredited, leading to social exclusion and rejection [7,8]. Infectious diseases such as TB and HIV are often associated with high levels of stigma, contributing to gaps in diagnosis, treatment, and care [6,9]. In the case of TB, stigma may arise from its association with HIV, fears of contagion, and misconceptions about its curability [7,10]. TB-related stigma may therefore be amplified by HIV coinfection, leading to overlapping forms of stigma in a phenomenon known as intersectional stigma, thereby worsening social exclusion and health disparities [9]. This intensified stigma can discourage individuals from seeking care and adhering to treatment, making it more difficult to control the spread of TB. For this paper, we focused on perceived TB-related stigma from the patient and community perspectives as well as internalized HIV-related stigma. Perceived TB-related stigma from patient perspective refers to how individuals with TB perceive the stigma directed to them whereas TB-related stigma from community perspective describes how members of the public perceive people with TB [11,12]. On the other hand, internalized HIV-related stigma describes when a person living with HIV accepts the negative societal attitudes, beliefs and stereotypes about HIV and applies it to themselves [13].The experiences of TB and HIV-related stigmas have been associated with poor mental health outcomes, particularly depression and anxiety which are highly prevalent among individuals with TB/HIV [14,15]. These mental health outcomes can act as significant barriers to effective TB treatment, leading to increased healthcare costs, poor treatment adherence and suboptimal treatment outcomes [15,16].

Meanwhile, gender plays an important role in TB and HIV-related stigma due to varying gender roles, expectations, inequalities, and sociocultural norms across genders [17,18]. As a result, males and females may experience disease-related stigma differently [17–19]. For example, studies have shown that whereas men express greater concern regarding the impact of TB stigma on their economic situations, women are often more worried about the negative effects on their marital prospects, which is in itself also linked to financial security for women in many societies [17,20,21]. Moreover, studies have shown that women are more likely to report higher intensity of shame and guilt with suicidal ideation and depression compared to men [17,22]. These findings suggest that there are prevailing differences regarding stigma experiences between males and females, which may influence the relationship between stigma and mental health challenges.

As a middle-income country, Botswana ranks among the nations with a high TB burden per capita, with an incidence rate of 244 cases per 100,000 population [2,3,23]. Understanding the role of gender in the relationship between TB/HIV-related stigma and mental health outcomes is essential, especially in settings like Botswana, where gender inequalities are well documented [17,24]. However, there is a scarcity of research examining the association between stigma and mental health outcomes among individuals living with TB/HIV and how gender may influence the association. Our previous study reported high prevalence of mental health outcomes among patients with TB in Botswana, as well as an association between mental health outcomes and household food insecurity [25]. Building upon these findings, this study aims to examine the association between TB/HIV-related stigma and mental health outcomes (depression and anxiety) and whether gender moderates the associations among people newly diagnosed with TB in Botswana.

## Methods

### Ethics Statement

Ethical approval for the study was obtained from the Institutional Review Boards (IRB) at the University of California, Irvine, and the Botswana Ministry of Health and Wellness, Health Research and Development Division. Potential participants were included in the study after assessing for eligibility and obtaining written informed consent through face-to-face interviews.

### Design and data collection

This current study analyzes data from a broader 2019 research project conducted across 12 health centers in Gaborone, Botswana. The original study was done to assess the association between mental health outcomes and a range of socio-behavioral factors among newly diagnosed TB patients. Our previous studies have analyzed different aspects of this dataset including the relationship between household food insecurity, HIV infection and mental health [25], depression and delayed TB treatment [16] and association between smoking and depression [22]. The current study focuses on the relationship between TB/HIV-related stigma and mental health outcomes, including gender moderation effects, which were not addressed in these earlier studies.

### Study population

People newly diagnosed with TB at 12 primary health centers who were 18 years or older were approached by research assistants after confirmation of TB diagnosis and review of their clinic registers. Potential participants were included after assessing for eligibility and obtaining written informed consent through face-to-face interviews. Interviews and questionnaires were administered in the local Setswana language or English by research assistants depending on the preference of the participant.

### Measures

In the original study, researcher-designed questionnaires were used to collect sociodemographic data of interest through face-to-face interviews. Sociodemographic data collected included gender, age, marital status, monthly income range, education, history of smoking and alcohol. Additional clinical information such as TB diagnosis, TB symptoms and HIV status were also collected from medical records. Standardized tools were utilized to assess food insecurity, TB-related stigma from patient perspective, TB-related stigma from community perspective, HIV-related stigma, depressive and anxiety symptoms [26–30].

In this present study, the Patient Health Questionnaire-9 (PHQ-9) was used to assess depressive symptoms. The PHQ-9 is composed of 9 items rated on the Likert scale and it is widely validated for use in primary health care settings in African regions, including Botswana [31–33]. The PHQ-9 achieved acceptable reliability among our study sample (Cronbach’s alpha = 0.74). The commonly used cutoff point of 10 was considered suggestive of significant symptoms of depression in the present study [16,27].

We assessed anxiety using the twenty-item ZUNG Self-Rating Anxiety Scale (ZUNG). The ZUNG is a self-report tool where respondents select how much each item applies to them according to a four-point Likert scale system [30,34]. The ZUNG assesses a range of neuro-cognitive, motor and autonomic symptoms of anxiety. A score of 36 or greater was considered indicative of anxiety symptoms [34,35]. Like the PHQ-9, the ZUNG achieved good internal reliability within our study population with Cronbach’s alpha of 0.80.

TB-related stigma from patient and community perspectives were measured using the perceived TB-related stigma scale developed by Van Rie and colleagues. The Perceived TB-related Stigma Scale (PTSS) has demonstrated good validity and reliability, achieving Cronbach’s alpha of 0.82 and 0.88 for patient and community perspectives, respectively [29]. Examples of questions from the PTSS from patient perspectives are “Some people who have TB lose friends when they share with them they have TB” and “Some people who have TB feel alone”. Similar items from the community perspective sub-scales are “Some people may not want to eat or drink with relatives who have TB” and “Some people prefer not to have those with TB living in their community”. Answer choices were rated on a Likert scale ranging from “strongly disagree”, “disagree”, “agree”, to “strongly agree” with higher scores indicating higher stigma. The PTSS achieved good internal reliability within our sample with Cronbach’s alpha of 0.89 and 0.93 for patient and community perspectives, respectively.

We measured HIV-related stigma using the ten-item HIV-related Internalized Stigma Scale (HISS) questionnaire [28]. HIV-related stigma was measured for only participants who tested positive for HIV. An example item from the HISS includes “How much do you feel that you have brought shame to your family because of your AIDS diagnosis”. Responses to the items were scored on a Likert system ranging from 0 (not at all) to 3 (great deal), with higher scores indicating higher stigma. The HISS demonstrated acceptable internal consistency achieving Cronbach’s alpha of 0.87.

Lastly, food insecurity was assessed using the nine-item Household Food Insecurity Access Scale (HFIAS), a widely used tool developed by the USAID Food and Nutrition Technical Assistance Project to estimate the level of food insecurity experienced within households [26,36]. An example item from the HFIAS is “In the past four weeks, did you or anyone in the house go to sleep at night hungry because there was not enough food?”. Responses ranged from 0 (Never) to 3 (Often). Higher HFIAS scores implied higher food insecurity within the household. The HFIAS achieved a Cronbach’s alpha of 0.94 in our sample.

### Statistical analysis

All data analyses were performed using R Statistical Software version 4.4.1. TB-related stigma from the patient and community perspectives was assessed using stigma scales developed by Van Rie and colleagues and were applied in the current study without modification [29].

Descriptive tables were used to summarize the variables from the study using mean and median values for continuous variables. Categorical variables were summarized using frequencies and percentages. We compared the HIV and TB-related stigma scores between males and females using Wilcoxon rank-sum tests. The Wilcoxon rank-sum test was used because the stigma scores showed non-uniform distribution after performing the Shapiro test. Results were presented using boxplots to show the median stigma scores and p-values for the Wilcoxon rank-sum test, using a statistically significant value of < 0.05.

We examined whether gender moderates the association between mental health outcomes and the three types of stigma (HIV-related stigma, TB-related stigma from the patient perspective, TB-related stigma from the community perspective) using linear regression models. Bivariate regression models were built to assess the association between each outcome variable and each type of stigma. Full linear multivariate regression models were then constructed for each association by including all selected covariates from our conceptual framework of the association between stigma types and mental health outcomes. Covariates that represent potential confounders included age, gender, marital status, TB-related stigma from the patient perspective, and TB-related stigma from the community perspective. Subsequently, new linear multivariate regression models were constructed to assess if the associations were moderated through gender by including the appropriate interaction terms (independent variable * gender) in the full multivariate models. Moderation analysis was done regardless of the results of the main effects model, as the main effects results may obscure associations with opposite directionality within moderator categories [37]. A P-value of < 0.1 for the interaction term was considered statistically significant. For this analysis, depression, anxiety, household food insecurity, HIV stigma score, and TB-related stigma were all analyzed as continuous variables. Sensitivity analysis was also done to assess whether the results would change based on the covariates that were included in our models. The first sensitivity analysis was done by excluding household food insecurity from the models since most similar studies did not include food insecurity. A second sensitivity analysis was done by including only TB-related stigma from patient and community perspectives in the models because several studies analyzed TB-related stigma from one perspective. Since the sample size for HIV positive individuals (N=99) may not have been sufficient for all the covariates fitted for the HIV-related stigma models, we did additional sensitivity analysis for HIV-related stigma by fitting models which included only covariates that were significant in the bivariate models.

## Results

Table 1 shows the characteristics of the study participants by gender. One hundred and eighty patients receiving TB treatment were included in the study, with 64 (35.6%) females and 99(55%) who tested HIV positive. The mean age of participants was 38.2 years, with the male participants being slightly older than females (39.5 years vs. 35.8 years). Majority of study participants (90%) were single and had less than secondary or some secondary school education (60.6%). Income levels differed, with 39.4% earning less than 800 Botswana Pula (59 US Dollars) per month; females were more likely to report earnings below 800 Pula (51.6% vs. 32.8%). Only 9.4% earned more than P5000 (370 US Dollars), with males being more likely to be within this earning bracket (12.9% vs. 3.1%).

**Table 1:** Characteristics of Study Participants by Gender.

|  | Female<br>(N=64) | Male<br>(N=116) | Overall<br>(N=180) |
| --- | --- | --- | --- |
| **Age** |  |  |  |
| Mean (SD) | 35.8 (12.1) | 39.5 (12.8) | 38.2 (12.6) |
| Median [Min, Max] | 33.5 [19.0, 76.0] | 39.0 [18.0, 68.0] | 38.0 [18.0, 76.0] |
| **Marital Status** |  |  |  |
| Single | 57 (89.1%) | 105 (90.5%) | 162 (90.0%) |
| Married | 7 (10.9%) | 11 (9.5%) | 18 (10.0%) |
| **Education Level** |  |  |  |
| Less than Secondary | 9 (14.1%) | 41 (35.3%) | 50 (27.8%) |
| Some Secondary | 26 (40.6%) | 33 (28.4%) | 59 (32.8%) |
| Completed Secondary | 14 (21.9%) | 17 (14.7%) | 31 (17.2%) |
| Post-Secondary | 15 (23.4%) | 25 (21.6%) | 40 (22.2%) |
| **Income (in Botswana pula\*)** |  |  |  |
| Less than P800 | 33 (51.6%) | 38 (32.8%) | 71 (39.4%) |
| P800–2999 | 22 (34.4%) | 40 (34.5%) | 62 (34.4%) |
| P3000–4999 | 7 (10.9%) | 23 (19.8%) | 30 (16.7%) |
| >P5000 | 2 (3.1%) | 15 (12.9%) | 17 (9.4%) |
| **TB Stigma Community Perspective** |  |  |  |
| Low stigma (0-11) | 11 (17.2%) | 19 (16.4%) | 30 (16.7%) |
| Moderate stigma (12-22) | 33 (51.6%) | 54 (46.6%) | 87 (48.3%) |
| High stigma (23-33) | 20 (31.3%) | 43 (37.1%) | 63 (35.0%) |
| **TB Stigma Patient Perspective** |  |  |  |
| Low stigma (0-12) | 8 (12.5%) | 13 (11.2%) | 21 (11.7%) |
| Moderate stigma (13-24) | 37 (57.8%) | 57 (49.1%) | 94 (52.2%) |
| High stigma (25-36) | 19 (29.7%) | 46 (39.7%) | 65 (36.1%) |
| **Household Food Insecurity Access Scale** |  |  |  |
| Secure | 33 (51.6%) | 63 (54.3%) | 96 (53.3%) |
| Mild insecure | 5 (7.8%) | 10 (8.6%) | 15 (8.3%) |
| Moderate insecure | 10 (15.6%) | 7 (6.0%) | 17 (9.4%) |
| Severe insecure | 16 (25.0%) | 36 (31.0%) | 52 (28.9%) |
| **HIV Status** |  |  |  |
| Negative | 27 (42.2%) | 54 (46.6%) | 81 (45.0%) |
| Positive | 37 (57.8%) | 62 (53.4%) | 99 (55.0%) |
| **HIV stigma score (HIV positive)** |  |  |  |
| Mean (SD) | 5.03 (6.35) | 5.72 (7.01) | 5.46 (6.74) |
| Median [Min, Max] | 2.00 [0, 19.0] | 3.00 [0, 24.0] | 3.00 [0, 24.0] |
| **Depression levels** |  |  |  |
| No depression (0) | 2 (3.1%) | 6 (5.2%) | 8 (4.4%) |
| Minimal depression (1-4) | 13 (20.3%) | 34 (29.3%) | 47 (26.1%) |
| Mild depression (5-9) | 28 (43.8%) | 50 (43.1%) | 78 (43.3%) |
| Moderate depression (10-14) | 14 (21.9%) | 12 (10.3%) | 26 (14.4%) |
| Severe depression (15-27) | 7 (10.9%) | 14 (12.1%) | 21 (11.7%) |
| **Anxiety levels** |  |  |  |
| Normal (20-35) | 31 (48.4%) | 64 (55.2%) | 95 (52.8%) |
| Minimal to moderate anxiety (36-47) | 27 (42.2%) | 44 (37.9%) | 71 (39.4%) |
| Marked to extreme anxiety ($\geq 48$) | 6 (9.4%) | 8 (6.9%) | 14 (7.8%) |

Over a third of participants reported high stigma for both community (35.0%) and patient (36.1%) TB-related stigma scales; 37.1 % of males and 31.3% of females reported high levels of stigma from the community perspective, whereas 39.7% of males and 29.7% of females reported high stigma levels from the patient perspective. The median community stigma score for male and female participants was 20 and 19.5, respectively (p = 0.32; Fig 1). The median score for TB-related stigma from the patient perspective for male and female participants was 22 and 21, respectively (p = 0.11; Fig 2). The mean HIV-related stigma score among HIV positive participants was 5.46, with male participants recording slightly higher mean scores than females (5.72 vs. 5.03, respectively). Median scores for HIV-related stigma was 3 for males and 2 for females (p = 0.67; Fig 3).

**Fig. 1.**
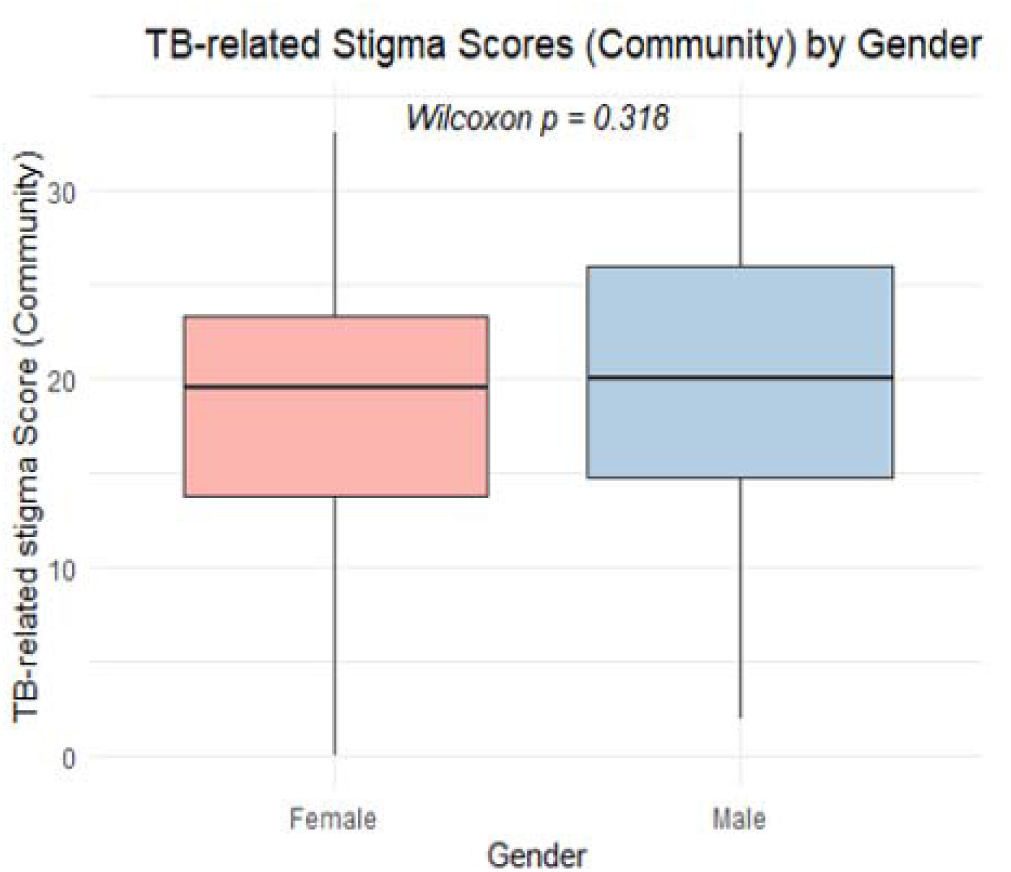

**Fig. 2.**
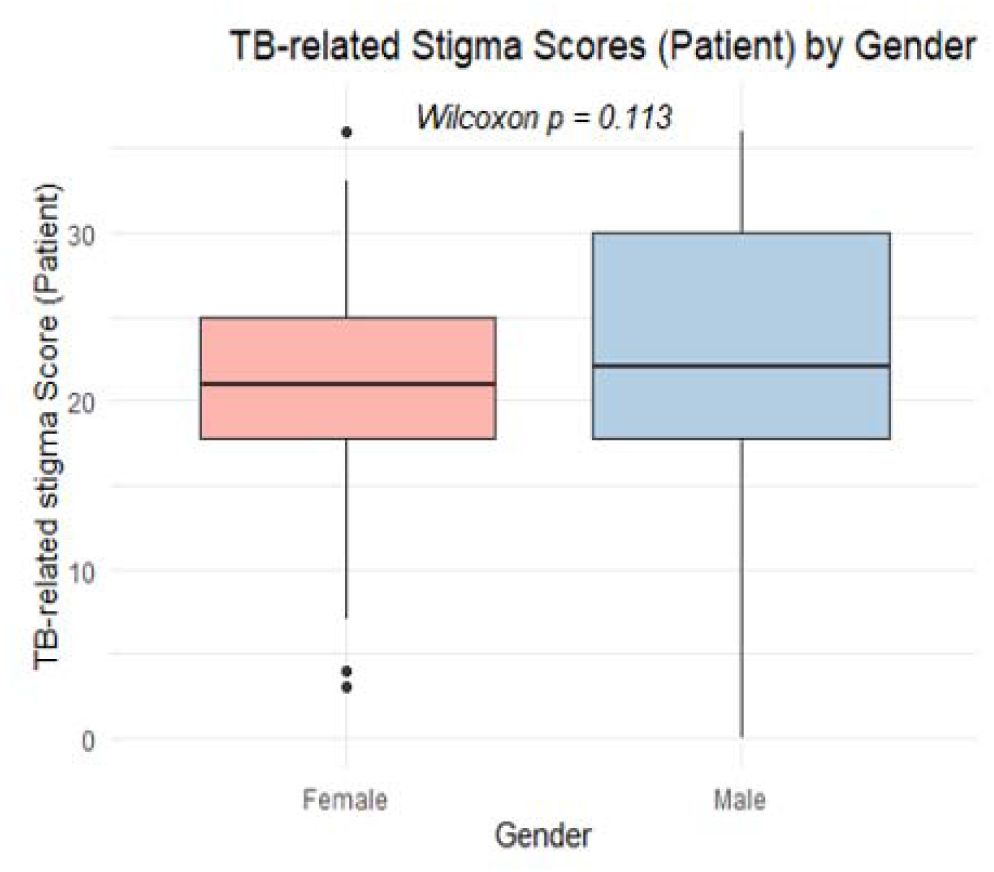

**Fig. 3.**
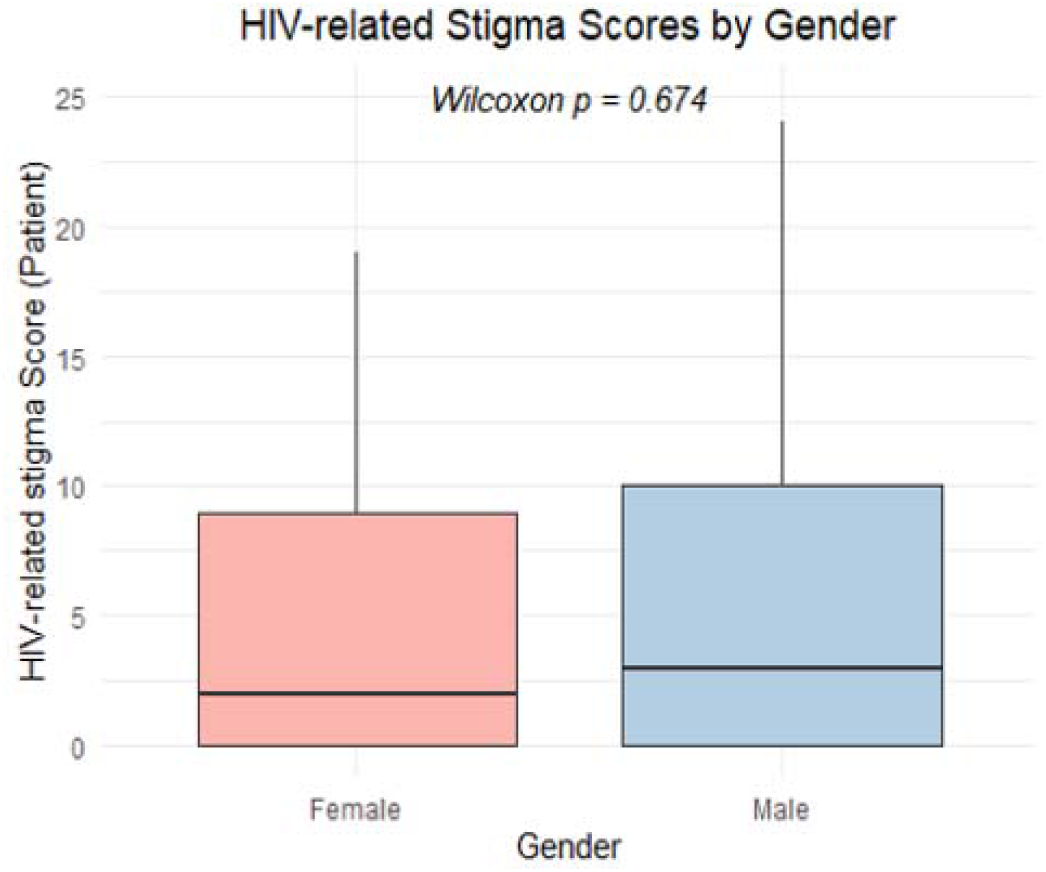

Ninety-six (53%) participants reported being food secure, 52(28.9%) were severely food insecure, while the remaining participants experienced mild or moderate food insecurity. Almost all participants (95.6%) reported some level of depression with 14.4% reporting moderate and 11.7% reporting severe depression. A higher proportion of males reported severe (12.1%) depression compared to female participants (10.9%), while female participants were more likely to report moderate depression (21.9% vs. 10.3%).

Just over half of the participants (52.8%) were categorized as having normal anxiety levels. Over one-third of participants (39.4%) reported minimal to moderate anxiety, while 7.8% experienced marked to most extreme anxiety. A slightly higher proportion of females than males reported both minimal to moderate anxiety (42.2% vs. 37.9%) and marked to most extreme anxiety (9.4% vs. 6.9%).

Table 2 presents the combined results for regression models examining the association between depression and TB-related stigmas (patient and community perspectives). Bivariate analysis showed that higher depression scores were associated with higher TB-related stigma from both patient (β = 0.17, p < 0.001) and community perspectives (β = 0.15, p = 0.002). When the other covariates were adjusted for, the associations between depression and TB-related stigma (patient) did not reach statistical significance (β = 0.12, p = 0.065). There was no evidence of association between depression and TB-related stigma (community) after adjusting for other covariates (β = 0.03, p = 0.7). In multivariable analysis, higher food insecurity was found to be associated with higher depression scores (β = 0.37, p < 0.001), and male participants were associated with lower depression scores (β = −2.0, p =0.015). Overall, there was no evidence of gender moderation in the associations between depression and TB-related stigma, with non-significant interaction terms for both patient (β = 0.04, p = 0.7) and community perspectives (β = 0.06, p = 0.5).

**Table 2:** Depression and TB-related Stigma model.

| Depression* | Bivariate |  | Multivariate |  | Moderation<br>(TB-stigma patient) |  | Moderation<br>(TB stigma community) |  |
| --- | --- | --- | --- | --- | --- | --- | --- | --- |
|  | Beta | p-value | Beta | p-value | Beta | p-value | Beta | p-value |
| **TB Stigma Patient Perspective** | 0.17 | <0.001 | 0.12 | 0.065 | 0.09 | 0.4 | 0.12 | 0.076 |
| **TB Stigma Community Perspective** | 0.15 | 0.002 | 0.03 | 0.7 | 0.03 | 0.6 | -0.01 | 0.934 |
| **Gender** |  |  |  |  |  |  |  |  |
| Female | — |  | — |  | — |  | — |  |
| Male | -0.93 | 0.3 | -2.0 | 0.015 | -2.9 | 0.2 | -3.2 | 0.11 |
| **Age** | 0.04 | 0.2 | -0.01 | 0.8 | -0.01 | 0.8 | -0.01 | 0.9 |
| **Marital Status** |  |  |  |  |  |  |  |  |
| Single | — |  | — |  | — |  | — |  |
| Married | -2.1 | 0.10 | -1.8 | 0.2 | -1.8 | 0.2 | -1.8 | 0.2 |
| **Education Level** |  |  |  |  |  |  |  |  |
| Less than Secondary | — |  | — |  | — |  | — |  |
| Some Secondary | -0.41 | 0.7 | -0.45 | 0.7 | -0.36 | 0.8 | -0.30 | 0.8 |
| Completed Secondary | -1.3 | 0.3 | -1.9 | 0.2 | -1.7 | 0.3 | -1.6 | 0.3 |
| Post Secondary | -2.8 | 0.012 | -2.9 | 0.059 | -2.8 | 0.077 | -2.7 | 0.094 |
| **Household Food Insecurity Access Scale** | 0.37 | <0.001 | 0.37 | <0.001 | 0.37 | <0.001 | 0.37 | <0.001 |
| **Income (Botswana pula\*)** |  |  |  |  |  |  |  |  |
| less than P800 | — |  | — |  | — |  | — |  |
| P800–2999 | -0.87 | 0.3 | -0.81 | 0.3 | -0.87 | 0.3 | -0.87 | 0.3 |
| P3000–4999 | -1.3 | 0.3 | 0.67 | 0.6 | 0.56 | 0.6 | 0.53 | 0.6 |
| >P5000 | -1.4 | 0.3 | -0.08 | 0.957 | -0.19 | 0.957 | -0.28 | 0.8 |
| **HIV Status** |  |  |  |  |  |  |  |  |
| Negative | — |  | — |  | — |  | — |  |
| Positive | 1.0 | 0.2 | -0.33 | 0.7 | -0.30 | 0.7 | -0.24 | 0.8 |
| **TB Stigma Patient Perspective \* Male** |  |  |  |  | 0.04 | 0.7 |  |  |
| **TB Stigma Community Perspective \* Male** |  |  |  |  |  |  | 0.06 | 0.5 |

The results for the regression models for anxiety and TB-related stigma (community and patient) are shown in Table 3. In the bivariate models, neither TB-related stigma from the patient perspective nor the community perspective showed any evidence of association with anxiety levels. The associations between anxiety and TB-related stigma (patient and community) did not reach statistical significance even after adjusting for the other covariates in the multivariate models (β = 0.04, p = 0.7 & β = −0.11, p = 0.4, respectively). Household food insecurity showed a strong positive association with increasing anxiety scores in the multivariate model (β = 0.55, p < 0.001). The multivariate model also demonstrated that participants who reported completing secondary education and those with post-secondary education had lower anxiety scores (β = −5.5, p = 0.045 & β = −7.5, p = 0.007) compared to participants with less than secondary education. Furthermore, male participants were also associated with lower anxiety scores (β = −3.7, p = 0.013) in the multivariate models. Overall, there was no evidence of moderation by gender for the associations between anxiety and TB-related stigma from the patient perspective (β = −0.18, p = 0.4) and community perspective (β = −0.12, p = 0.5).

**Table 3:** Anxiety and TB-related Stigma Model.

| Anxiety* | Bivariate |  | Multivariate |  | Moderation<br>(TB-stigma patient) |  | Moderation<br>(TB-stigma community) |  |
| --- | --- | --- | --- | --- | --- | --- | --- | --- |
|  | Beta | p-value | Beta | p-value | Beta | p-value | Beta | p-value |
| **TB Stigma Patient Perspective** | 0.04 | 0.6 | 0.04 | 0.7 | 0.18 | 0.4 | 0.05 | 0.7 |
| **TB Stigma Community Perspective** | 0.03 | 0.8 | -0.11 | 0.4 | -0.12 | 0.3 | -0.04 | 0.8 |
| **Gender** |  |  |  |  |  |  |  |  |
| Female | — |  | — |  | — |  | — |  |
| Male | -2.6 | 0.072 | -3.7 | 0.013 | 0.04 | 0.9925 | -1.5 | 0.7 |
| **Age** | 0.02 | 0.8 | -0.11 | 0.2 | -0.11 | 0.2 | -0.11 | 0.2 |
| **Marital Status** |  |  |  |  |  |  |  |  |
| Single | — |  | — |  | — |  | — |  |
| Married | -1.2 | 0.6 | 0.05 | 0.983 | 0.17 | 0.944 | 0.12 | 0.959 |
| **Education Level** |  |  |  |  |  |  |  |  |
| Less than Secondary | — |  | — |  | — |  | — |  |
| Some Secondary | 0.58 | 0.7 | -1.7 | 0.4 | -2.0 | 0.4 | -1.9 | 0.4 |
| Completed Secondary | -1.9 | 0.4 | -5.5 | 0.045 | -6.0 | 0.032 | -5.9 | 0.036 |
| Post Secondary | -3.6 | 0.065 | -7.5 | 0.007 | -8.0 | 0.005 | -7.9 | 0.006 |
| **Household Food Insecurity Access Scale** | 0.49 | <0.001 | 0.55 | <0.001 | 0.56 | <0.001 | 0.57 | <0.001 |
| **Income (Botswana pula\*)** |  |  |  |  |  |  |  |  |
| less than P800 | — |  | — |  | — |  | — |  |
| P800–2999 | -2.8 | 0.081 | -2.8 | 0.071 | -2.6 | 0.11 | -2.7 | 0.086 |
| P3000–4999 | -3.0 | 0.13 | 0.16 | 0.938 | 0.64 | 0.8 | 0.43 | 0.8 |
| >P5000 | -1.4 | 0.6 | 2.0 | 0.4 | 2.5 | 0.3 | 2.4 | 0.4 |
| **HIV Status** |  |  |  |  |  |  |  |  |
| Negative | — |  | — |  | — |  | — |  |
| Positive | 2.6 | 0.058 | 1.3 | 0.4 | 1.2 | 0.4 | 1.1 | 0.4 |
| **TB Stigma Patient Perspective \* Male** |  |  |  |  | -0.18 | 0.4 |  |  |
| **TB Stigma Community Perspective \* Male** |  |  |  |  |  |  | -0.12 | 0.5 |

Table 4 presents the results for the models (bivariate, multivariate, and moderation) fitted for the association between depression and HIV-related stigma. In the bivariate model, higher HIV-related stigma scores were associated with increased depression scores (β = 0.17, p = 0.033). After adjusting for age, gender, educational level, income, marital status, household food insecurity, and TB-related stigma, the association did not reach statistical significance (β = 0.07, p = 0.4). In the multivariate model, food insecurity and male gender showed association with increasing and decreasing depression scores, respectively (β = 0.38, p < 0.001 vs β = −3.0, p = 0.013). The interaction between HIV-related stigma and gender was not significant (β = −0.06, p = 0.7), indicating that gender did not moderate the relationship between HIV-related stigma and depression.

**Table 4:** Depression and HIV-related Stigma Model.

| Depression* | Bivariate |  | Multivariate |  | Moderation |  |
| --- | --- | --- | --- | --- | --- | --- |
|  | Beta | p-value | Beta | p-value | Beta | p-value |
| **HIV-related stigma score** | 0.17 | 0.033 | 0.07 | 0.4 | 0.11 | 0.4 |
| **Gender** |  |  |  |  |  |  |
| Female | — |  | — |  | — |  |
| Male | -0.93 | 0.3 | -3.0 | 0.013 | -2.7 | 0.069 |
| **Age** | 0.04 | 0.2 | -0.03 | 0.7 | -0.02 | 0.7 |
| **Marital Status** |  |  |  |  |  |  |
| Single | — |  | — |  | — |  |
| Married | -2.1 | 0.10 | -3.2 | 0.086 | -3.1 | 0.10 |
| **Education Level** |  |  |  |  |  |  |
| Less than Secondary | — |  | — |  | — |  |
| Some Secondary | -0.41 | 0.7 | -2.3 | 0.13 | -2.3 | 0.14 |
| Completed Secondary | -1.3 | 0.3 | -2.6 | 0.2 | -2.6 | 0.2 |
| Post-Secondary | -2.8 | 0.012 | -2.9 | 0.2 | -3.0 | 0.2 |
| **Household Food Insecurity Access Scale** | 0.37 | <0.001 | 0.38 | <0.001 | 0.39 | <0.001 |
| **Income (Botswana pula\*)** |  |  |  |  |  |  |
| Less than P800 | — |  | — |  | — |  |
| P800–2999 | -0.87 | 0.3 | -1.8 | 0.2 | -1.8 | 0.2 |
| P3000–4999 | -1.3 | 0.3 | 2.6 | 0.10 | 2.6 | 0.10 |
| >P5000 | -1.4 | 0.3 | 0.18 | 0.926 | 0.17 | 0.929 |
| **TB Stigma Patient Perspective** | 0.17 | <0.001 | -0.02 | 0.8 | -0.03 | 0.8 |
| **TB Stigma Community Perspective** | 0.15 | 0.002 | 0.11 | 0.3 | 0.12 | 0.3 |
| **HIV-related stigma score \* Male** |  |  |  |  | -0.06 | 0.7 |

Table 5 shows the results for the regression models examining the association between anxiety and HIV-related Stigma. Overall, the results showed that the association between anxiety and HIV-related stigma was not moderated by gender (β = −0.27, p = 0.4). In the bivariate model, higher HIV-related stigma was associated with higher anxiety scores (β = 0.31, p = 0.038). This association was, however, reduced and no longer reached statistical significance in the multivariate model (β = 0.24, p = 0.14). Male gender was associated with lower anxiety scores in the multivariate model (β = –5.6, p = 0.018), whilst higher food insecurity was associated with higher anxiety scores (β = 0.54, p = 0.007).

**Table 5:** Anxiety and HIV-related Stigma Model.

| Anxiety* | Bivariate |  | Multivariate |  | Moderation |  |
| --- | --- | --- | --- | --- | --- | --- |
|  | Beta | p-value | Beta | p-value | Beta | p-value |
| **HIV-related stigma score** | 0.31 | 0.038 | 0.24 | 0.14 | 0.42 | 0.12 |
| **Gender** |  |  |  |  |  |  |
| Female | — |  | — |  | — |  |
| Male | -2.6 | 0.072 | -5.6 | 0.018 | -4.2 | 0.15 |
| **Age** | 0.02 | 0.8 | -0.15 | 0.3 | -0.14 | 0.3 |
| **Marital Status** |  |  |  |  |  |  |
| Single | — |  | — |  | — |  |
| Married | -1.2 | 0.6 | 1.4 | 0.7 | 1.7 | 0.6 |
| **Education Level** |  |  |  |  |  |  |
| Less than Secondary | — |  | — |  | — |  |
| Some Secondary | 0.58 | 0.7 | -2.9 | 0.3 | -2.7 | 0.4 |
| Completed Secondary | -1.9 | 0.4 | -5.0 | 0.2 | -5.1 | 0.2 |
| Post -Secondary | -3.6 | 0.065 | -7.9 | 0.061 | -8.1 | 0.055 |
| **Income (Botswana pula\*)** |  |  |  |  |  |  |
| Less than P800 | — |  | — |  | — |  |
| P800–2999 | -2.8 | 0.081 | -2.6 | 0.3 | -2.5 | 0.3 |
| P3000–4999 | -3.0 | 0.13 | 1.8 | 0.6 | 1.9 | 0.5 |
| >P5000 | -1.4 | 0.6 | 4.7 | 0.2 | 4.7 | 0.2 |
| **Household Food Insecurity Access Scale** | 0.49 | <0.001 | 0.54 | 0.007 | 0.56 | 0.006 |
| **TB Stigma Patient Perspective** | 0.04 | 0.6 | -0.02 | 0.901 | -0.04 | 0.8 |
| **TB Stigma Community Perspective** | 0.03 | 0.8 | -0.09 | 0.7 | -0.06 | 0.8 |
| **HIV-related stigma score \* Male** |  |  |  |  | -0.27 | 0.4 |

Overall, the results of our sensitivity analysis were consistent with the primary analyses. Excluding household food insecurity from the TB-related stigma models did not change the associations, confirming that our findings were robust. Likewise, the sensitivity model including only the TB-related stigma variables without adjustment for any other covariates yielded similar results, indicating that the primary models were not over-adjusted. However, when household food insecurity was excluded from the HIV-related stigma model, an association between HIV-related stigma and anxiety was observed. Sensitivity analysis results are presented in **S**upplementary tables (S1-8 Tables).

## Discussion

### Association between TB-related stigma and mental health outcomes

The main objectives of our study were to explore the association between TB/HIV-related stigma and mental health outcomes and determine whether gender acts as a moderator for those associations. The results from our study showed that TB-related stigma from the patient and community perspectives were not associated with mental health outcomes (depression and anxiety symptoms) in the final multivariate models. The findings of this current study contradict those of a multi-site cross-sectional survey of 612 Indonesian adults with TB which found that TB-related stigma (patient and community) was positively associated with depression [38]. Likewise, a study by Assefa and his colleagues found that TB patients who reported perceived stigma were three times more likely to experience anxiety [39].

There may be several reasons why the results of this current study contradict other previous studies. Firstly, the observed difference could be because of the covariates included in our study. For instance, the inclusion of TB-related stigma from both patient and community perspectives as covariates may have impacted the results. In the initial bivariate models, both TB-related stigmas showed a positive association with depression. This association remained after adjusting for sociodemographic covariates, HIV status, and food insecurity. The association disappeared only when TB-related stigma (either patient or community) was included in the model, indicating that TB-related stigma (patient) or TB stigma (community) had a substantial diminishing effect on the other in the models. However, there was no evidence of multicollinearity upon assessment using the variance inflation factor. Additionally, contextual factors in Botswana may be different from studies conducted in other settings such as Ethiopia, China, and Indonesia. Botswana is one of the few countries that has achieved the UNAIDS 95-95-95 goal, which aims for 95% of people living with HIV to know their status, 95% of those diagnosed to be on treatment, and 95% of those on treatment to achieve viral suppression by 2025 [40,41]. This significant progress in TB/HIV integrated care may have reduced the effect of TB and or HIV-related stigma on mental health outcomes in our sample as compared to other settings where studies have reported heightened fear and stigma attached to the disease [38,39,42].

### Association between HIV-related stigma and mental health outcomes

Contrary to previous studies, we did not find any association between HIV-related stigma and mental health outcomes in our sample. The pooled estimates of Ethiopian studies exploring the relationship between HIV-related stigma and depression revealed a significant association between HIV stigma and depression [43]. The meta-analysis showed that participants with perceived stigma were twice as likely to experience depression [43]. Our finding also contradicts several other studies that have reported significant associations between HIV stigma and mental health outcomes [44–47]. The bivariate models showed association between HIV-related stigma and mental health outcomes which disappeared after adjusting for other covariates in the main effects models. HIV-related stigma was associated with anxiety when food insecurity was excluded from the model suggesting that food insecurity may have been the main confounder for the observed association between HIV-related stigma and anxiety (S4 Table).

Consistent with previous studies, food insecurity was consistently associated with mental health outcomes in our sample reinforcing its negative impact on mental health among people with TB/HIV in Botswana [48–50]. It is possible that mental health outcomes in our study population may be driven by food insecurity and its related socioeconomic hardships rather than infection-related stigma [51,52].

### Gender as a moderator

Our results showed that gender did not moderate the associations between TB and or HIV-related stigma and mental health outcomes. We also found comparable median TB and or HIV-related stigma scores between men and women, suggesting that both genders may experience stigma at comparable levels in our sample. Prior research has examined gender as a moderator in the relationship between TB stigma and other outcomes such as treatment acceptance [53]. To the best of our knowledge, this is the first study examining gender as a moderator for the association between TB/HIV-related stigma and mental health outcomes among TB patients. Future studies should be conducted to explore the gender moderating effects for the TB/HIV stigma - mental health outcome link in different settings where stigma levels are significantly different across gender.

#### Strengths and limitations

This study is one of the few studies that has attempted to explain the role of gender as a moderator for the TB or HIV-related stigma and mental health outcomes link. We also acknowledge that our study has several limitations. Firstly, our study is a cross-sectional study, which limits its ability to draw causal inferences. Additionally, our results may not be generalizable to other settings, because all participants were recruited from health centers in Gaborone. This present study also did not measure variables such as social support and coping strategies that may moderate this relationship. Finally, there is the possibility of self-report bias in our study because the measurement tools relied on participants’ answers.

## Conclusion

Contrary to prior research, we found no significant association between either TB or HIV-related stigma and mental health outcomes. Moreover, our study suggests that gender does not moderate the association between TB and or HIV-related stigma and mental health outcomes. We recommend conducting further studies with larger sample sizes to explore the nature of the relationship between mental health outcomes and TB or HIV-related stigma among patients with TB. Additionally, these studies should investigate the potential of other related factors such as social support and coping strategies as moderators for this relationship. Notably, food insecurity and gender were found to be consistently associated with mental health outcomes. This finding supports the recommendation for TB programs to integrate strategies that address food insecurity. We further recommend that these TB programs should be strengthened and extended to address food insecurity as well as other related financial and material needs of people with TB and or HIV. Gender specific strategies may be needed to address the more severe mental health outcomes among women.

## Data Availability

All data produced in the present study are available upon reasonable request to the authors

https://doi.org/10.5281/zenodo.17259915

## Acknowledgement

We would like to express gratitude to all participants and research staff who devoted their time and efforts make this study possible. We are also thankful to the University of California Irvine and Botswana Ministry of Health and Wellness Human Research Development Division. We are also grateful to all the research staff for their efforts. The authors declare no conflicts of interest.

