## Supplemental Table 1 for "Gender and the associations between HIV and TB-related stigma and mental health outcomes among people with tuberculosis in Botswana"

|  | **Bivariate** | | **Multivariate** | | **Moderation**  **(TB-stigma patient)** | | **Moderation**  **(TB-stigma community)** | |
| --- | --- | --- | --- | --- | --- | --- | --- | --- |
| Depression* | **Beta** | **p-value** | **Beta** | **p-value** | **Beta** | **p-value** | **Beta** | **p-value** |
| **TB Stigma Patient Perspective** | 0.17 | <0.001 | 0.11 | 0.11 | 0.04 | 0.8 | 0.10 | 0.15 |
| **TB Stigma Community Perspective** | 0.15 | 0.002 | 0.06 | 0.4 | 0.06 | 0.4 | -0.03 | 0.7 |
| **Gender** |  |  |  |  |  |  |  |  |
| Female | — |  | — |  | — |  | — |  |
| Male | -0.93 | 0.3 | -1.4 | 0.12 | -3.5 | 0.2 | -4.2 | 0.049 |
| **Age** | 0.04 | 0.2 | 0.00 | 0.949 | 0.01 | 0.906 | 0.01 | 0.9 |
| **Marital Status** |  |  |  |  |  |  |  |  |
| Single | — |  | — |  | — |  | — |  |
| Married | -2.1 | 0.10 | -2.4 | 0.088 | -2.5 | 0.082 | -2.5 | 0.080 |
| **Education Level** |  |  |  |  |  |  |  |  |
| Less than Secondary | — |  | — |  | — |  | — |  |
| Some Secondary | -0.41 | 0.7 | -0.31 | 0.8 | -0.11 | 0.935 | 0.03 | 0.983 |
| Completed Secondary | -1.3 | 0.3 | -1.1 | 0.5 | -0.86 | 0.6 | -0.65 | 0.7 |
| Post-Secondary | -2.8 | 0.012 | -2.3 | 0.2 | -2.0 | 0.2 | -1.7 | 0.3 |
| **Income (Botswana pula*)** |  |  |  |  |  |  |  |  |
| less than P800 | — |  | — |  | — |  | — |  |
| P800–2999 | -0.87 | 0.3 | -0.91 | 0.3 | -1.1 | 0.3 | -1.1 | 0.3 |
| P3000–4999 | -1.3 | 0.3 | -0.30 | 0.8 | -0.55 | 0.7 | -0.59 | 0.6 |
| >P5000 | -1.4 | 0.3 | -0.62 | 0.7 | -0.88 | 0.6 | -1.1 | 0.5 |
| **HIV Status** |  |  |  |  |  |  |  |  |
| Negative | — |  | — |  | — |  | — |  |
| Positive | 1.0 | 0.2 | -0.04 | 0.962 | 0.02 | 0.980 | 0.15 | 0.9 |
| **TB Stigma Patient Perspective * Male** |  |  |  |  | 0.10 | 0.4 |  |  |
| **TB Stigma Community Perspective * Male** |  |  |  |  |  |  | 0.15 | 0.15 |
