## Supplemental Table 2 for "Gender and the associations between HIV and TB-related stigma and mental health outcomes among people with tuberculosis in Botswana"

|  | **Bivariate** | | **Multivariate** | | **Moderation**  **(TB-stigma patient)** | | **Moderation**  **(TB stigma community)** | |
| --- | --- | --- | --- | --- | --- | --- | --- | --- |
| **Anxiety*** | **Beta** | **p-value** | **Beta** | **p-value** | **Beta** | **p-value** | **Beta** | **p-value** |
| **TB Stigma Patient Perspective** | 0.04 | 0.6 | 0.03 | 0.8 | 0.09 | 0.6 | 0.02 | 0.8 |
| **TB Stigma Community Perspective** | 0.03 | 0.8 | -0.07 | 0.6 | -0.07 | 0.6 | -0.08 | 0.6 |
| **Gender** |  |  |  |  |  |  |  |  |
| Female | — |  | — |  | — |  | — |  |
| Male | -2.6 | 0.072 | -2.7 | 0.075 | -0.87 | 0.8 | -3.1 | 0.4 |
| **Age** | 0.02 | 0.8 | -0.09 | 0.3 | -0.09 | 0.3 | -0.09 | 0.3 |
| **Marital Status** |  |  |  |  |  |  |  |  |
| Single | — |  | — |  | — |  | — |  |
| Married | -1.2 | 0.6 | -0.86 | 0.7 | -0.81 | 0.7 | -0.87 | 0.7 |
| **Education Level** |  |  |  |  |  |  |  |  |
| Less than Secondary | — |  | — |  | — |  | — |  |
| Some Secondary | 0.58 | 0.7 | -1.5 | 0.5 | -1.6 | 0.5 | -1.4 | 0.5 |
| Completed Secondary | -1.9 | 0.4 | -4.4 | 0.12 | -4.6 | 0.11 | -4.4 | 0.14 |
| Post-secondary | -3.6 | 0.065 | -6.5 | 0.025 | -6.8 | 0.023 | -6.5 | 0.031 |
| **Income (Botswana pula*)** |  |  |  |  |  |  |  |  |
| Less than P800 | — |  | — |  | — |  | — |  |
| P800–2999 | -2.8 | 0.081 | -3.0 | 0.070 | -2.8 | 0.089 | -3.0 | 0.071 |
| P3000–4999 | -3.0 | 0.13 | -1.3 | 0.5 | -1.0 | 0.6 | -1.3 | 0.5 |
| >P5000 | -1.4 | 0.6 | 1.2 | 0.7 | 1.4 | 0.6 | 1.1 | 0.7 |
| **HIV Status** |  |  |  |  |  |  |  |  |
| Negative | — |  | — |  | — |  | — |  |
| Positive | 2.6 | 0.058 | 1.7 | 0.2 | 1.7 | 0.3 | 1.7 | 0.3 |
| **TB Stigma Patient Perspective * Male** |  |  |  |  | -0.09 | 0.7 |  |  |
| **TB Stigma Community Perspective * Male** |  |  |  |  |  |  | 0.02 | 0.922 |
