## Supplemental Table 4 for "Gender and the associations between HIV and TB-related stigma and mental health outcomes among people with tuberculosis in Botswana"

|  | **Bivariate** | | **Multivariate** | | **Moderation** | |
| --- | --- | --- | --- | --- | --- | --- |
| Anxiety* | **Beta** | **p-value** | **Beta** | **p-value** | **Beta** | **p-value** |
| **HIV-related stigma score** | 0.31 | 0.038 | 0.35 | 0.037 | 0.49 | 0.085 |
| **Gender** |  |  |  |  |  |  |
| Female | — |  | — |  | — |  |
| Male | -2.6 | 0.072 | -4.2 | 0.080 | -3.1 | 0.3 |
| **Age** | 0.02 | 0.8 | -0.12 | 0.4 | -0.11 | 0.4 |
| **Marital Status** |  |  |  |  |  |  |
| Single | — |  | — |  | — |  |
| Married | -1.2 | 0.6 | 0.88 | 0.8 | 1.1 | 0.8 |
| **Education Level** |  |  |  |  |  |  |
| Less than Secondary | — |  | — |  | — |  |
| Some Secondary | 0.58 | 0.7 | -1.9 | 0.5 | -1.8 | 0.6 |
| Completed Secondary | -1.9 | 0.4 | -3.0 | 0.5 | -2.9 | 0.5 |
| Post-Secondary | -3.6 | 0.065 | -6.7 | 0.12 | -6.8 | 0.12 |
| **Income (Botswana pula*)** |  |  |  |  |  |  |
| less than P800 | — |  | — |  | — |  |
| P800–2999 | -2.8 | 0.081 | -3.0 | 0.2 | -3.0 | 0.3 |
| P3000–4999 | -3.0 | 0.13 | -0.11 | 0.971 | -0.04 | 0.989 |
| >P5000 | -1.4 | 0.6 | 3.3 | 0.4 | 3.2 | 0.4 |
| **TB Stigma Patient Perspective** | 0.04 | 0.6 | -0.04 | 0.9 | -0.05 | 0.8 |
| **TB Stigma Community Perspective** | 0.03 | 0.8 | -0.06 | 0.8 | -0.04 | 0.9 |
| **HIV-related stigma score * Male** |  |  |  |  | -0.20 | 0.5 |
