## Supplemental Table 5 for "Gender and the associations between HIV and TB-related stigma and mental health outcomes among people with tuberculosis in Botswana"

| Depression* | **Beta** | **95% CI** | **p-value** |
| --- | --- | --- | --- |
| **HIV-related stigma score** | 0.07 | -0.10, 0.24 | 0.4 |
| **Education Level** |  |  |  |
| Less than Secondary | — | — |  |
| Some Secondary | -0.49 | -3.0, 2.0 | 0.7 |
| Completed Secondary | -0.49 | -3.6, 2.6 | 0.8 |
| Post Secondary | -0.64 | -4.0, 2.7 | 0.7 |
| **TB Stigma Community Perspective** | 0.10 | -0.10, 0.30 | 0.3 |
| **TB Stigma Patient Perspective** | 0.00 | -0.19, 0.20 | >0.9 |
| **Household Food Insecurity Access Scale** | 0.32 | 0.11, 0.52 | 0.002 |
| Abbreviation: CI = Confidence Interval  This model was fitted using only covariates that had significant bivariate association  in Table 4. | | | |
