## Supplemental Table 6 for "Gender and the associations between HIV and TB-related stigma and mental health outcomes among people with tuberculosis in Botswana"

| Anxiety* | **Beta** | **95% CI** | **p-value** |
| --- | --- | --- | --- |
| **HIV-related stigma score** | 0.21 | -0.09, 0.50 | 0.2 |
| **Household Food Insecurity Access Scale** | 0.42 | 0.05, 0.79 | 0.026 |
| Abbreviation: CI = Confidence Interval  This model was fitted using only covariates that had significant bivariate association  in Table 5. | | | |
