## Supplemental Table 7 for "Gender and the associations between HIV and TB-related stigma and mental health outcomes among people with tuberculosis in Botswana"

| Depression* | **Beta** | **95% CI** | **p-value** |
| --- | --- | --- | --- |
| TB Stigma Community Perspective | 0.08 | -0.05, 0.21 | 0.2 |
| TB Stigma Patient Perspective | 0.12 | -0.02, 0.25 | 0.085 |
| Abbreviation: CI = Confidence Interval  This model was fitted with only TB-related stigma from community and patient perspectives | | | |
