## Supplemental Table 8 for "Gender and the associations between HIV and TB-related stigma and mental health outcomes among people with tuberculosis in Botswana"

| Anxiety* | **Beta** | **95% CI** | **p-value** |
| --- | --- | --- | --- |
| TB Stigma Community Perspective | 0.00 | -0.23, 0.23 | 0.978 |
| TB Stigma Patient Perspective | 0.05 | -0.19, 0.28 | 0.7 |
| Abbreviation: CI = Confidence Interval  This model was fitted with only TB-related stigma from community and patient perspectives | | | |
